# A Systematic Review of Machine Learning-based Prognostic Models for Acute Pancreatitis: Towards Improving Methods and Reporting Quality

**DOI:** 10.1101/2024.06.26.24309389

**Authors:** Brian Critelli, Amier Hassan, Ila Lahooti, Lydia Noh, Jun Sung Park, Kathleen Tong, Ali Lahooti, Nate Matzko, Jan Niklas Adams, Lukas Liss, Justin Quion, David Restrepo, Melica Nikahd, Stacey Culp, Adam Lacy-Hulbert, Cate Speake, James Buxbaum, Jason Bischof, Cemal Yazici, Anna Evans Phillips, Sophie Terp, Alexandra Weissman, Darwin Conwell, Phil Hart, Mitch Ramsey, Somashekar Krishna, Samuel Han, Erica Park, Raj Shah, Venkata Akshintala, John A Windsor, Nikhil K Mull, Georgios I Papachristou, Leo Anthony Celi, Peter J Lee

**Affiliations:** Weill Cornell Medical College, Division of Gastroenterology and Hepatology; Ohio State University Wexner Medical Center, Division of Gastroenterology and Hepatology; Northeast Ohio Medical School; Rheinisch-Westfälische Technische Hochschule Aachen University, Division of Process and Data Science; Massachusetts Institute of Technology, Laboratory for Computational Physiology; Ohio State University Wexner Medical Center, Division of Bioinformatics; Center for Systems Immunology, Benaroya Research Institute at Virginia Mason, Seattle, Washington; Center for Interventional Immunology, Benaroya Research Institute at Virginia Mason, Seattle, Washington; University of Southern California, Division of Gastroenterology; Ohio State University Wexner Medical Center, Division of Emergency Medicine; University of Illinois at Chicago, Division of Gastroenterology; University of Pittsburgh Medical Center, Division of Gastroenterology; University of Southern California, Division of Emergency Medicine; University of Pittsburgh Medical Center, Division of Emergency Medicine; University of Kentucky, Department of Medicine; Johns Hopkins Medical Center, Division of Gastroenterology; University of Auckland, Surgical and Translational Research Centre; University of Pennsylvania, Division of Hospital Medicine and Penn Medicine Center for Evidence-based Practice; Beth Israel Medical Center, Division of Critical Care

## Abstract

**Background:** An accurate prognostic tool is essential to aid clinical decision making (e.g., patient triage) and to advance personalized medicine. However, such prognostic tool is lacking for acute pancreatitis (AP). Increasingly machine learning (ML) techniques are being used to develop high-performing prognostic models in AP. However, **methodologic and reporting quality has received little attention**. High-quality reporting and study methodology are critical to model validity, reproducibility, and clinical implementation. In collaboration with content experts in ML methodology, we performed a systematic review critically appraising the quality of methodology and reporting of recently published ML AP prognostic models.

**Methods:** Using a validated search strategy, we identified ML AP studies from the databases MEDLINE, PubMed, and EMBASE published between January 2021 and December 2023. Eligibility criteria included all retrospective or prospective studies that developed or validated new or existing ML models in patients with AP that predicted an outcome following an episode of AP. Meta-analysis was considered if there was homogeneity in the study design and in the type of outcome predicted. For risk of bias (ROB) assessment, we used the Prediction Model Risk of Bias Assessment Tool (PROBAST). Quality of reporting was assessed using the Transparent Reporting of a Multivariable Prediction Model of Individual Prognosis or Diagnosis – Artificial Intelligence (TRIPOD+AI) statement that defines standards for 27 items that should be reported in publications using ML prognostic models.

**Results:** The search strategy identified 6480 publications of which 30 met the eligibility criteria. Studies originated from China (22), U.S (4), and other (4). All 30 studies developed a new ML model and none sought to validate an existing ML model, producing a total of 39 new ML models. AP severity (23/39) or mortality (6/39) were the most common outcomes predicted. The mean area-under-the-curve for all models and endpoints was 0.91 (SD 0.08). The ROB was high for at least one domain in all 39 models, particularly for the analysis domain (37/39 models). Steps were not taken to minimize over-optimistic model performance in 27/39 models. Due to heterogeneity in the study design and in how the outcomes were defined and determined, meta-analysis was not performed.

Studies reported on only 15/27 items from TRIPOD+AI standards, with only 7/30 justifying sample size and 13/30 assessing data quality. Other reporting deficiencies included omissions regarding human-AI interaction (28/30), handling low-quality or incomplete data in practice (27/30), sharing analytical codes (25/30), study protocols (25/30) and reporting source data (19/30),.

**Discussion:** There are significant deficiencies in the methodology and reporting of recently published ML based prognostic models in AP patients. These undermine the validity, reproducibility and implementation of these prognostic models despite their promise of superior predictive accuracy.

**Funding:** none

**Registration:** Research Registry ([reviewregistry1727](https://www.researchregistry.com/browse-the-registry#registryofsystematicreviewsmeta-analyses/registryofsystematicreviewsmeta-analysesdetails/653bcd876d91ab002791fee6/))

## INTRODUCTION

Defined as acute inflammation of the pancreas, acute pancreatitis (AP) remains a common and costly cause of gastrointestinal-related hospitalization, with 1 million new cases each year globally and increasing incidence[1, 2]. The etiology of the disease varies across patient demographics, with gallstones and alcohol comprising the majority of adult cases and diverse environmental factors such as hypertriglyceridemia, drugs, infections, or trauma[3]. The severity of AP can be further categorized as mild, moderately severe, or severe, with severe AP being defined by the presence of persistent organ failure [4]. The combination of persistent organ failure and infected pancreatic necrosis defines a ‘critical’ category of AP severity with the highest morbidity and mortality risk[5, 6]. Survivors of AP can suffer from long-term sequelae including diabetes mellitus, recurrent or chronic pancreatitis, and pancreatic exocrine insufficiency[3, 7–10]. Given the significant short- and long-term morbidity and mortality associated with AP, the National Institute of Health has called for an accurate prognostic model in AP for use in research and the clinical setting[11–13]. Benefits of an accurate prognostic model are many, including enablement of cost-efficient clinical trials through cohort enrichment [14, 15], identification of subphenotypes within a cohort that require different treatment strategies [16, 17], and prompt triaging of patients in the emergency room [18].

Current prognostic models for AP were developed using regression-based techniques (e.g., Glasgow Criteria, Bedside Index for Severity in Acute Pancreatitis (BISAP) etc.) which demonstrate suboptimal performance and limited clinical usefulness[19]. For example, in a prospective external evaluation of regression-based models predicting mortality, none of the models tested produced a post-test probability higher than 14% when “positive”[20]. There has been a call for new approaches to improve prediction accuracy [19, 21]. Advances in the subset of artificial intelligence (AI) known as machine learning (ML) have facilitated the development of non-regression prediction models, which offer advantages over regression-based models by performing better in diseases with non-linear predictor-outcome relationships such as AP[22]. There has been an increasing number of published ML-based prognostic models that appear to outperform regression-based models [23–25]. However, ML experts have cited concerns regarding methodologic quality, model building practices, and lack of transparent reporting [26–28]. We therefore undertook a systematic review and critical appraisal of recent published studies proposing new non-regression ML based prognostic models to detail any methodological shortcomings and/or gaps in reporting. This was a collaborative effort between experienced clinicians and ML experts [19].

## METHODS

Detailed methodology of this review has been published elsewhere[29] (doi: 10.1186/s41512-024-00169-1). We conducted a systematic review of all studies published between January 2021 and December 2023 in which a non-regression, ML-based prognostic model in AP was developed and/or validated (either internally or externally), with or without model updating. This review included studies of prospective or retrospective design including post-hoc analysis of clinical trials that: a) enrolled only adult patients (i.e., 18 years old or older), b) contained a prognostic model of AP developed with non-regression ML technique(s), c) predicted any outcome(s) of AP, and d) published in English. Studies involving participants with chronic pancreatitis, pancreatic cancer, or post-surgical pancreatitis were excluded, as were studies with animals, regression-based models, or models that predict the development of AP instead of disease outcomes. Studies published in abstract form only and review articles were also excluded.

We searched the databases MEDLINE (OvidSP) and EMBASE (OvidSP) from January 1, 2021 to December 31, 2023 (Date of search for all data sources, January 31^st^) Our search was limited to the most recent three years for the following reasons 1) Significant advancements in AP management paradigm has led to a significant change in the natural history/prognosis of the disease over the last decade [30–37]. It was important to identify models trained/evaluated on datasets generated from the most recent cohort of AP. 2) New algorithms rapidly emerge, replacing older algorithms and temporal quality degradation is an established phenomenon in AI models[38]. Validated search strategies [39, 40] were used and are listed in Supplementary Tables 1 and 2, respectively. Covidence software (city, country) was used to screened title-abstract and full text in sequential steps. Each stage required concordance between two independent reviewers (LN, IL, KT, JP, AH, BC, NM, or AL). Disagreements were resolved by a third independent reviewer (PJL or LAC). Included studies were then appraised in terms of risk of bias in study design, completeness of reporting, and for summarization of model predictive performances. Necessary data for PROBAST and TRIPOD+AI evaluation were extracted in accordance with the Critical Appraisal and Data Extraction for Systematic Reviews of Prediction Modelling Studies (CHARMS) checklist[41].

### Methodologic Quality Assessment

The Prediction Model Risk of Bias Assessment Tool (PROBAST) was used to assess both risk of bias in study design of prospective models across four main domains: participants, predictors, outcomes, and analysis[42]. The assessment of Applicability section of PROBAST was planned if meta-data were appropriate and feasible for meta-analysis. To optimize the validity of the PROBAST assessment, all evaluators underwent PROBAST rater training, which entailed weekly meetings with an AP content expert trained by PROBAST developers (PJL) to review all 20 signaling questions. Data scientists (JNA, LL, JQ, or DR) and ML content experts (LAC) were engaged to accurately complete CHARMS and PROBAST. Each model was assessed via the PROBAST framework by two independent reviewers (LN, IL, KT, JP, AH, BC, NM, AL, JNA, LL, JQ, or DR), and disagreements were resolved by an independent third reviewer (PJL or LAC). The pair of reviewers comprised a clinician and a data scientist. The risk of bias in each domain and overall risk of bias were reported for all studies.

### Reporting Quality Assessment

To assess the quality of the reporting, we decided to use TRIPOD+AI statement, which contains a comprehensive list of items that need to be reported for papers reporting development and/or validation of prognostic AI model[43]. List of sections and items on this list covers every key part of a manuscript including title, abstract, introduction, methods, results, and discussion. Additionally, it contains items related to open science and patient & public involvement. Summary statistics of quality of reporting according to the standards of TRIPOD+AI[43] were calculated for each study. This review has been registered at Research Registry ([reviewregistry1727](https://www.researchregistry.com/browse-the-registry#registryofsystematicreviewsmeta-analyses/registryofsystematicreviewsmeta-analysesdetails/653bcd876d91ab002791fee6/)). All data reporting in this systematic review adhered to the guidelines of Preferred Reporting Items for Systematic reviews and Meta-Analyses (PRISMA) and the checklist can be found in a separate supplementary file.

## RESULTS

Metadata used to generate these results can be accessed at DOI:10.6084/m9.figshare.26078743. Our search strategy identified 6480 studies published between January 2021 and December 2023, of which 30 met eligibility criteria (S1 Figure). Studies originated from China (22), the United States (4), Hungary (2), Turkey (1), and New Zealand (1) (Table 1).

**Table 1:**
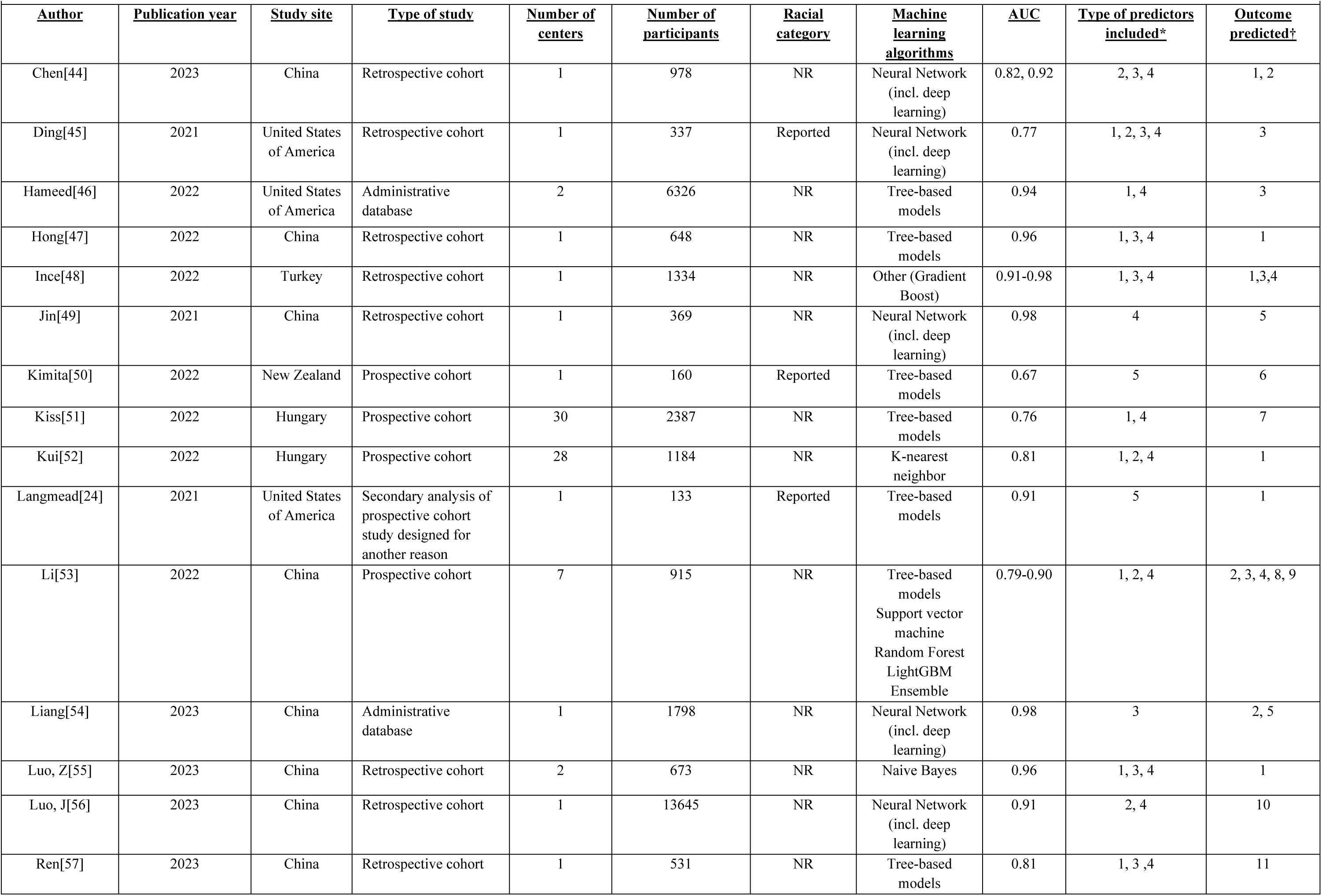
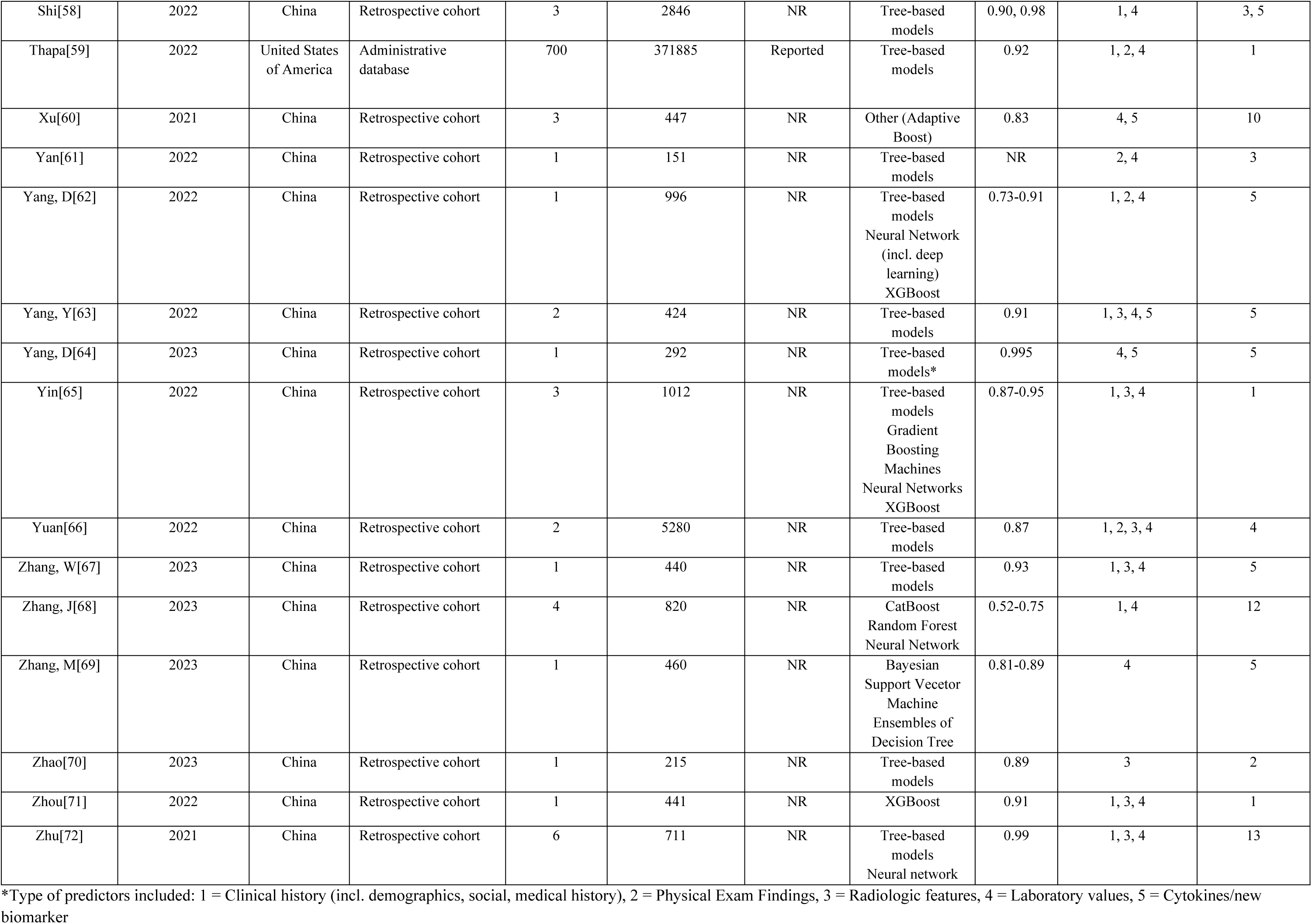
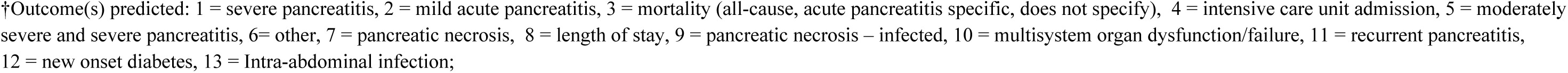
Basic characteristics of included studies.

| <u>Author</u> | <u>Publication year</u> | <u>Study site</u> | <u>Type of study</u> | <u>Number of centers</u> | <u>Number of participants</u> | <u>Racial category</u> | <u>Machine learning algorithms</u> | <u>AUC</u> | <u>Type of predictors included*</u> | <u>Outcome predicted†</u> |
| --- | --- | --- | --- | --- | --- | --- | --- | --- | --- | --- |
| Chen[44] | 2023 | China | Retrospective cohort | 1 | 978 | NR | Neural Network (incl. deep learning) | 0.82, 0.92 | 2, 3, 4 | 1, 2 |
| Ding[45] | 2021 | United States of America | Retrospective cohort | 1 | 337 | Reported | Neural Network (incl. deep learning) | 0.77 | 1, 2, 3, 4 | 3 |
| Hameed[46] | 2022 | United States of America | Administrative database | 2 | 6326 | NR | Tree-based models | 0.94 | 1, 4 | 3 |
| Hong[47] | 2022 | China | Retrospective cohort | 1 | 648 | NR | Tree-based models | 0.96 | 1, 3, 4 | 1 |
| Ince[48] | 2022 | Turkey | Retrospective cohort | 1 | 1334 | NR | Other (Gradient Boost) | 0.91-0.98 | 1, 3, 4 | 1,3,4 |
| Jin[49] | 2021 | China | Retrospective cohort | 1 | 369 | NR | Neural Network (incl. deep learning) | 0.98 | 4 | 5 |
| Kimita[50] | 2022 | New Zealand | Prospective cohort | 1 | 160 | Reported | Tree-based models | 0.67 | 5 | 6 |
| Kiss[51] | 2022 | Hungary | Prospective cohort | 30 | 2387 | NR | Tree-based models | 0.76 | 1, 4 | 7 |
| Kui[52] | 2022 | Hungary | Prospective cohort | 28 | 1184 | NR | K-nearest neighbor | 0.81 | 1, 2, 4 | 1 |
| Langmead[24] | 2021 | United States of America | Secondary analysis of prospective cohort study designed for another reason | 1 | 133 | Reported | Tree-based models | 0.91 | 5 | 1 |
| Li[53] | 2022 | China | Prospective cohort | 7 | 915 | NR | Tree-based models<br>Support vector machine<br>Random Forest<br>LightGBM<br>Ensemble | 0.79-0.90 | 1, 2, 4 | 2, 3, 4, 8, 9 |
| Liang[54] | 2023 | China | Administrative database | 1 | 1798 | NR | Neural Network (incl. deep learning) | 0.98 | 3 | 2, 5 |
| Luo, Z[55] | 2023 | China | Retrospective cohort | 2 | 673 | NR | Naive Bayes | 0.96 | 1, 3, 4 | 1 |
| Luo, J[56] | 2023 | China | Retrospective cohort | 1 | 13645 | NR | Neural Network (incl. deep learning) | 0.91 | 2, 4 | 10 |
| Ren[57] | 2023 | China | Retrospective cohort | 1 | 531 | NR | Tree-based models | 0.81 | 1, 3, 4 | 11 |
| Shi[58] | 2022 | China | Retrospective cohort | 3 | 2846 | NR | Tree-based models | 0.90, 0.98 | 1, 4 | 3, 5 |
| Thapa[59] | 2022 | United States of America | Administrative database | 700 | 371885 | Reported | Tree-based models | 0.92 | 1, 2, 4 | 1 |
| Xu[60] | 2021 | China | Retrospective cohort | 3 | 447 | NR | Other (Adaptive Boost) | 0.83 | 4, 5 | 10 |
| Yan[61] | 2022 | China | Retrospective cohort | 1 | 151 | NR | Tree-based models | NR | 2, 4 | 3 |
| Yang, D[62] | 2022 | China | Retrospective cohort | 1 | 996 | NR | Tree-based models<br>Neural Network (incl. deep learning)<br>XGBoost | 0.73-0.91 | 1, 2, 4 | 5 |
| Yang, Y[63] | 2022 | China | Retrospective cohort | 2 | 424 | NR | Tree-based models | 0.91 | 1, 3, 4, 5 | 5 |
| Yang, D[64] | 2023 | China | Retrospective cohort | 1 | 292 | NR | Tree-based models* | 0.995 | 4, 5 | 5 |
| Yin[65] | 2022 | China | Retrospective cohort | 3 | 1012 | NR | Tree-based models<br>Gradient Boosting Machines<br>Neural Networks<br>XGBoost | 0.87-0.95 | 1, 3, 4 | 1 |
| Yuan[66] | 2022 | China | Retrospective cohort | 2 | 5280 | NR | Tree-based models | 0.87 | 1, 2, 3, 4 | 4 |
| Zhang, W[67] | 2023 | China | Retrospective cohort | 1 | 440 | NR | Tree-based models | 0.93 | 1, 3, 4 | 5 |
| Zhang, J[68] | 2023 | China | Retrospective cohort | 4 | 820 | NR | CatBoost<br>Random Forest<br>Neural Network | 0.52-0.75 | 1, 4 | 12 |
| Zhang, M[69] | 2023 | China | Retrospective cohort | 1 | 460 | NR | Bayesian<br>Support Vecetor Machine<br>Ensembles of Decision Tree | 0.81-0.89 | 4 | 5 |
| Zhao[70] | 2023 | China | Retrospective cohort | 1 | 215 | NR | Tree-based models | 0.89 | 3 | 2 |
| Zhou[71] | 2022 | China | Retrospective cohort | 1 | 441 | NR | XGBoost | 0.91 | 1, 3, 4 | 1 |
| Zhu[72] | 2021 | China | Retrospective cohort | 6 | 711 | NR | Tree-based models<br>Neural network | 0.99 | 1, 3, 4 | 13 |

\*Type of predictors included: 1 = Clinical history (incl. demographics, social, medical history), 2 = Physical Exam Findings, 3 = Radiologic features, 4 = Laboratory values, 5 = Cytokines/new biomarker
†Outcome(s) predicted: 1 = severe pancreatitis, 2 = mild acute pancreatitis, 3 = mortality (all-cause, acute pancreatitis specific, does not specify), 4 = intensive care unit admission, 5 = moderately severe and severe pancreatitis, 6= other, 7 = pancreatic necrosis, 8 = length of stay, 9 = pancreatic necrosis – infected, 10 = multisystem organ dysfunction/failure, 11 = recurrent pancreatitis, 12 = new onset diabetes, 13 = Intra-abdominal infection;

All 30 studies reported the development of a new ML-based prognostic model, but only one study included external validation step of the newly developed model. Nearly three-fourths (22/30) of included studies were retrospective cohort, while only five studies were prospective, of which one was a secondary analysis. Five studies developed more than one model, resulting in a total of 39 models developed in 30 studies. The most common machine learning algorithms were tree-based models (20/39) and neural networks (7/39). AP severity (21/39) or mortality (6/39) were the most common outcomes predicted. The most common methods of internal validation were cross-validation (23/39) and bootstrapping (17/39). For 31/39 models, shrinkage methods were not used to evaluate for or adjust for optimism (shrinkage methods: techniques used to account for magnitude of noise in the dataset contributing to overinflation of predictive performance). A summary of pertinent descriptive statistics collected as per the CHARMS checklist is provided in Table 1. Overall, for the 39 models the mean area-under-the-curve (AUC) was 0.91 (SD 0.08). Six studies developed more than one ML-model using the same dataset, presenting the parameters of the “best performing” model (Table 1). Every model had at least one domain in which the risk of bias was classified as high (Fig 1), meaning that all 39 models were assessed to be at high risk of bias by PROBAST standards (see S3 Table for individual model’s ROB rating). The median number of TRIPOD+AI items that were reported on in the 30 studies was 15/27 (range 6-20). No study reported on all the items. A comprehensive breakdown of the number of TRIPOD+AI items reported on in each study is given in Supplementary Table 4 and on the heatmap for visual presentation of the data (Fig 2).

**Fig 1.**
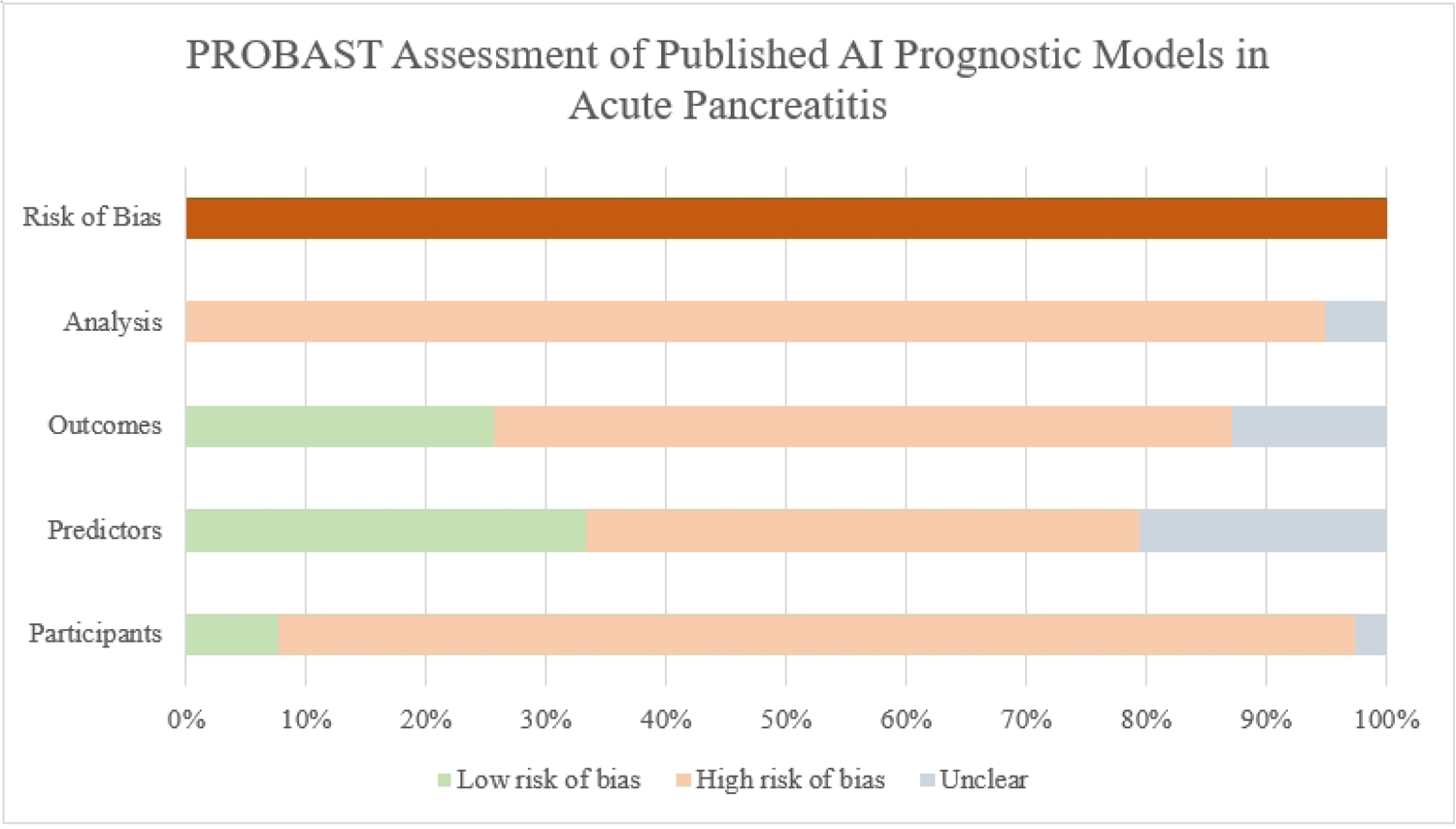
Summary of Risk of Bias in 4 Domains Assessed by PROBAST.

**Fig 2.**
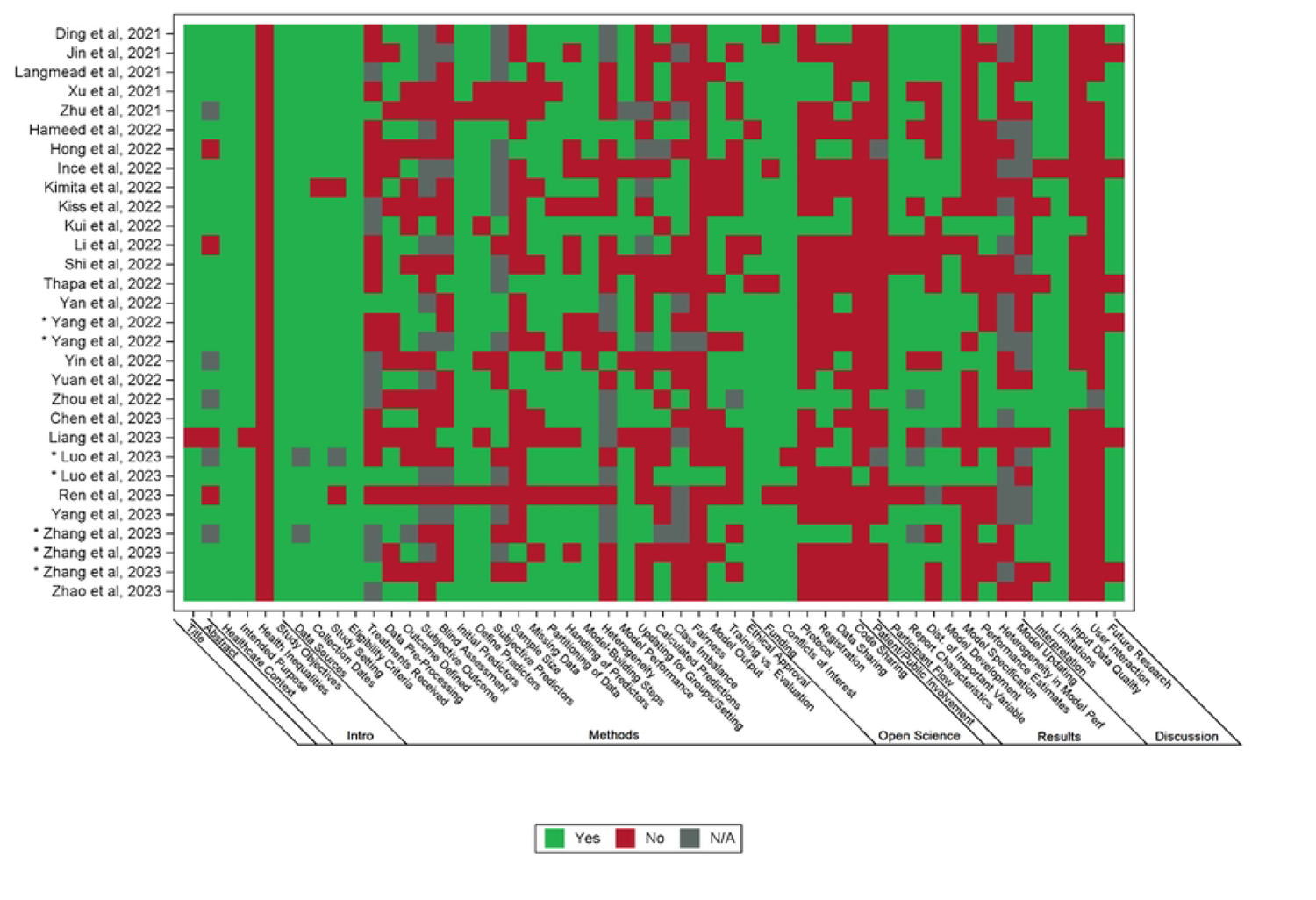
Heatmap depicting common areas of deficiencies in reporting standards as assessed by TRIPOD+AI. *Publication has same first author and year as another paper listed; PMID of each * in ascending order: Yang et al, 2022: 35430680, 35607360. Luo et al, 2023: 36653317, 36773821. Zhang et al, 2023: 36902504, 36964219, 37196588.

### Risk of Bias in Four Domains of Methodology as Assessed by PROBAST

PROBAST ratings of the 39 models based on individual studies are summarized in Supplementary Table 3. Assessment of Applicability was not applicable to the objectives of this review. As the primary objective was to assess the methodologic quality and because of marked heterogeneity of the cohorts and the different definitions and determination of outcomes, a synthesis of the meta-data was not undertaken.

#### Participants Domain

In this domain there was a high risk of bias with 35/39 models. The data source was not appropriate with 31/39 models. The inclusions and exclusions of participants was not appropriate in 26/39 models.

#### Predictors Domain

In this domain there was a high risk of bias with 18/39 models. The predictors were not defined and measured in a similar way for all participants in 12/39 models. Assessor blinding to the outcome data was not done with 30/39 models. In 8/39 studies predictors were included when the result would not be available at the time of applying the prognostic model.

#### Outcomes Domain

In this domain there was a high risk of bias with 24/39 models. While outcomes were *defined* in a standard way in 33/39 models, they were not *determined* appropriately in 20/39 models. The way that outcomes were determined was not reported for 1/39 models. Outcomes were not defined and determined in a similar way in 13/39 models. Blinding was not performed in 24/39 models. Outcomes were included as predictors in 17/39 models.

#### Analysis Domain

In this domain there was a high risk of bias with 37/39 models (Fig 1). The common deficiencies in this domain were no accounting for overfitting and optimism (i.e. no shrinkage methods employed) in 31/39 models, none or inappropriate reporting of data complexity in 38/39 models (Fig 2), insufficient sample size in 28/39 models, and selection of predictors relied solely on univariate analysis in 26/39 models.

### Quality of Reporting as Assessed by TRIPOD+AI

#### Title, Abstract, Introduction Section

All 30 studies reported to the standards of TRIPOD+AI except in one important sub-item. No study reported the health inequalities that may exist in outcomes between sociodemographic groups (Fig 2 and S4 Table).

#### Methods Section

Twenty-eight studies described The sources of data, study dates, setting and eligibility were described in 28/30 studies but only 5/30 studies reported details of any treatment received where treatment might have influences the outcome of interest. Other frequent omissions included no description of model fairness and their rationale (28/30), no sample size justification (23/30), no blinding of assessors (20/30), no reporting differences between training and evaluation data (16/30), no outcome measurement (15/30), no description of data preparation and pre-processing(13/30), no reporting of elements pertinent to outcome definition (13/30), and no assessment of study quality (13/30)

#### Open Science and Patient/Public Involvement Section

There was no reporting on whether a protocol was prepared, available or accessed in 25/30 studies. There was no report as to the availability of study data (9/30) or analytical code (28/30). There was comment on whether patients and public were involved in 26/30 studies.

#### Results Section

There was insufficient detail of the prediction model to allow external validation in 25/30 studies. Reporting details of the prediction model performance in key subgroups (e.g. sociodemographic) was not available in 15/30 studies.

#### Discussion Section

Items pertaining to the usability of the model in the context of current care were usually not discussed. Only 3/30 studies described how poor quality or missing data should be handled with clinical implementation of the model. Only 1/30 study specified whether users will be required to interact with handling of the input data or use of the model and what level of expertise is required to use the model.

#### Rationale against performing subgroup analyses

Even though several of the included studies developed models predicting similar outcomes, decision was made not to perform subgroup analyses stratified by similar endpoints. All but one model was judged to have high risk of bias in at least two out of the four PROBAST domains and none of the models were at low risk of bias in the statistical analyses domain. With such limitations in the methodology across the board, subgroup analyses were felt not to lead to meaningful discoveries or different conclusions.

## DISCUSSION

In this systematic review, we assessed the quality of the methodology and reporting of studies that develop and/or validated non-regression ML-based models in AP literature. While the performance of the published models was high (mean AUC 0.91), we identified several key limitations in the recently published models. Unfortunately, these shortcomings are like those identified in other fields such as oncology[28] and anesthesiology[73]. First, the concern relates to the high risk of bias most notably in the statistical analysis section, which can undermine the validity of the models. Second, due to the lack of external validation studies, generalizability of the ML models may be limited. Third relates to open science practice, where in over 90% of the studies, the code was not shared and no information was provided on how the model was built. Additionally, there was a lack of reporting on how the ML model can be implemented in clinical practice. Lastly, none of the studies described potential health inequities among different sociodemographic groups, which risks widening disparities in healthcare, if implemented in real clinical practice.

The quality of the statistical analyses is one of the most important facets of model development. The PROBAST ROB tool dedicates 9 signaling questions to this domain[42]. Two particularly deficient areas were sample size justification and guarding against overfitting. A robust sample size (especially for a ML model) and guarding against overfitting are critically important. When these steps are omitted, a model may perform well in the development dataset but the predictive performance may not be reproducible[74]. We found that most published studies developed a model with a sample size of less than 1,000 participants and median events per variable was 9.5. Even for regression-based models, the minimum recommended events per variable is 20[42]. While events per variable is not a singular reflection of sufficient sample size, it is generally accepted that ML models require much larger sample size (than regression-based models) due to the risk of model instability[75].

Potentially limited generalizability of the published models need to be highlighted. Only one study conducted external validation but with limitations, all but 5 studies were single-center design. While AP is a common gastrointestinal disease, with an annual worldwide 1 million new cases a year[76], international or large multi-center consortiums with efforts to build a generalizable model have been lacking. Lack of such collaboration results in siloed attempts at building models that may not be clinically utilized due to poor reproducibility and generalizability. As with the case with the regression-based models[21], we are seeing a similar trend in ML-based models in AP.

Ultimately, prognostic models are built to aid clinical decision making or enhance cohort enrichment in a research study. Therefore, steps need to be taken to thoughtfully consider real-life issues we will face when trying to deploy these models (e.g., ways to deal with missing values in real clinical practice when patients won’t have the data elements necessary for the ML model). We also found key missing items relevant to open science, that limit external validation studies by other investigators and clinical implementation by the hospitals. For example, only 5 studies shared the code to permit third-party evaluation and implementation, only 3 studies gave guidance on how to handle missing data, and one study detailed the specifics of what constitutes human-AI interaction. As important, aspects of model building relevant healthcare equity (e.g., comparison of performance estimates among different sociodemographic subgroups) were not evaluated. Such deficiency leads to a potential to produce a model that widens the socioeconomic disparities[77].

Our study has several strengths. For transparency and rigor of our methodology we have published our methods and adhered strictly to the standards of TRIPOD-SR/MA. Our work was conducted in collaboration between data scientists, ML methodologist, and content experts in AP, which we believe enhances the reliability of our findings. There are multiple aspects to PROBAST and TRIPOD+AI assessment that require both AP content and ML methodology expertise. Third, rigorous internal training for PROBAST assessment preceded the project, enhancing the validity of our ROS assessment.

Several limitations deserve mention. Our search strategy extended only the last 3 years so it is possible that our findings may not be fully representative of all the ML models published for AP thus far. Second, while PROBAST was developed by expert methodologists, it is possible that models deemed high ROB by PROBAST may still be valid, reproducible, and generalizable in AP. However, there is emerging data from other diseases that suggest models deemed high ROB by PROBAST perform poorly external validation studies[78, 79]

In conclusion, the potential benefit of ML-based prognostic models is evident with an overall high AUC (mean 0.91±0.8SD). However, this study indicates that there should be great caution in implementing the reported models because of the very major concerns with the quality of the methodology and reporting. These raise questions about the validity, reproducibility, and generalizability of the prognostic models. It is recommended that AP-specific, standardized methodology that covers all 4 PROBAST domains and all items within TRIPOD+AI be used in developing and validating ML-based prognostic models. Only then should implementation be considered. Our study findings provide valuable baseline assessment of the quality of methods and reporting of ML-based models in AP. It is also timely given the recent publication of TRIPOD+AI[43], which we hope will encourage future investigators to utilize.

## Supporting information

Prisma diagrams

## Data Availability

https://figshare.com/s/64a07bd4eb2a0f334e69 DOI: 10.6084/m9.figshare.26078743

https://doi.org/10.6084/m9.figshare.26078743

## ACKNOWLEDGEMENTS

none

## REFERENCES

1. Xiao AY, Tan ML, Wu LM, Asrani VM, Windsor JA, Yadav D, Petrov MS. Global incidence and mortality of pancreatic diseases: a systematic review, meta-analysis, and meta-regression of population-based cohort studies. Lancet Gastroenterol Hepatol. 2016;1(1):45–55. Epub 20160628. doi: 10.1016/s2468-1253(16)30004-8. PubMed PMID: 28404111.

2. Iannuzzi JP, King JA, Leong JH, Quan J, Windsor JW, Tanyingoh D, et al. Global Incidence of Acute Pancreatitis Is Increasing Over Time: A Systematic Review and Meta-Analysis. Gastroenterology. 2022;162(1):122–34. Epub 20210925. doi: 10.1053/j.gastro.2021.09.043. PubMed PMID: 34571026.

3. Lee PJ, Papachristou GI. New insights into acute pancreatitis. Nature reviews Gastroenterology & hepatology. 2019. doi: 10.1038/s41575-019-0158-2.

4. Banks PA, Bollen TL, Dervenis C, Gooszen HG, Johnson CD, Sarr MG, et al. Classification of acute pancreatitis--2012: revision of the Atlanta classification and definitions by international consensus. Gut. 2013;62(1):102–11. doi: 10.1136/gutjnl-2012-302779 [doi].

5. Dellinger EP, Forsmark CE, Layer P, Levy P, Maravi-Poma E, Petrov MS, et al. Determinant-based classification of acute pancreatitis severity: an international multidisciplinary consultation. Annals of surgery. 2012;256(6):875–80. doi: 10.1097/SLA.0b013e318256f778.

6. Wu D, Lu B, Xue H-D, Yang H, Qian J-M, Lee P, Windsor JA. Validation of Modified Determinant-Based Classification of severity for acute pancreatitis in a tertiary teaching hospital. Pancreatology : official journal of the International Association of Pancreatology (IAP) [et al]. 2019;19(2):217–23. doi: 10.1016/j.pan.2019.01.003.

7. Petrov MS, Yadav D. Global epidemiology and holistic prevention of pancreatitis. Nat Rev Gastroenterol Hepatol. 2019;16(3):175–84. doi: 10.1038/s41575-018-0087-5. PubMed PMID: 30482911; PubMed Central PMCID: PMCPMC6597260.

8. Das SL, Singh PP, Phillips AR, Murphy R, Windsor JA, Petrov MS. Newly diagnosed diabetes mellitus after acute pancreatitis: a systematic review and meta-analysis. Gut. 2014;63(5):818–31. doi: 10.1136/gutjnl-2013-305062 [doi].

9. Huang W, de la Iglesia-García D, Baston-Rey I, Calviño-Suarez C, Lariño-Noia J, Iglesias-Garcia J, et al. Exocrine Pancreatic Insufficiency Following Acute Pancreatitis: Systematic Review and Meta-Analysis. Digestive diseases and sciences. 2019;64(7):1985–2005. doi: 10.1007/s10620-019-05568-9.

10. Zhi M, Zhu X, Lugea A, Waldron RT, Pandol SJ, Li L. Incidence of New Onset Diabetes Mellitus Secondary to Acute Pancreatitis: A Systematic Review and Meta-Analysis. Front Physiol. 2019;10:637. Epub 20190531. doi: 10.3389/fphys.2019.00637. PubMed PMID: 31231233; PubMed Central PMCID: PMCPMC6558372.

11. Abu-El-Haija M, Gukovskaya AS, Andersen DK, Gardner TB, Hegyi P, Pandol SJ, et al. Accelerating the Drug Delivery Pipeline for Acute and Chronic Pancreatitis: Summary of the Working Group on Drug Development and Trials in Acute Pancreatitis at the National Institute of Diabetes and Digestive and Kidney Diseases Workshop. Pancreas2018. p. 1185–92.

12. Uc A, Andersen DK, Borowitz D, Glesby MJ, Mayerle J, Sutton R, Pandol SJ. Accelerating the Drug Delivery Pipeline for Acute and Chronic Pancreatitis-Knowledge Gaps and Research Opportunities: Overview Summary of a National Institute of Diabetes and Digestive and Kidney Diseases Workshop. Pancreas2018. p. 1180–4.

13. Serrano J, Laughlin MR, Bellin MD, Yadav D, Chinchilli VM, Andersen DK. Type 1 Diabetes in Acute Pancreatitis Consortium: From Concept to Reality. Pancreas. 2022;51(6):563–7. doi: 10.1097/mpa.0000000000002073. PubMed PMID: 36206459; PubMed Central PMCID: PMCPMC9555854.

14. van Brunschot S, van Grinsven J, Voermans RP, Bakker OJ, Besselink MG, Boermeester MA, et al. Transluminal endoscopic step-up approach versus minimally invasive surgical step-up approach in patients with infected necrotising pancreatitis (TENSION trial): design and rationale of a randomised controlled multicenter trial [ISRCTN09186711. BMC gastroenterology. 2013;13:161-. doi: 10.1186/1471-230X-13-161 [doi].

15. van Santvoort HC, Besselink MG, Bakker OJ, Hofker HS, Boermeester MA, Dejong CH, et al. A step-up approach or open necrosectomy for necrotizing pancreatitis. The New England journal of medicine. 2010;362(16):1491–502. doi: 10.1056/NEJMoa0908821 [doi].

16. Giamarellos-Bourboulis EJ, Aschenbrenner AC, Bauer M, Bock C, Calandra T, Gat-Viks I, et al. The pathophysiology of sepsis and precision-medicine-based immunotherapy. Nature Immunology. 2024;25(1):19–28. doi: 10.1038/s41590-023-01660-5.

17. Rosenson RS, Gaudet D, Ballantyne CM, Baum SJ, Bergeron J, Kershaw EE, et al. Evinacumab in severe hypertriglyceridemia with or without lipoprotein lipase pathway mutations: a phase 2 randomized trial. Nat Med. 2023;29(3):729–37. Epub 20230306. doi: 10.1038/s41591-023-02222-w. PubMed PMID: 36879129; PubMed Central PMCID: PMCPMC10033404.

18. Raita Y, Goto T, Faridi MK, Brown DFM, Camargo CA, Jr., Hasegawa K. Emergency department triage prediction of clinical outcomes using machine learning models. Critical care (London, England). 2019;23(1):64-. doi: 10.1186/s13054-019-2351-7.

19. Capurso G, Ponz de Leon Pisani R, Lauri G, Archibugi L, Hegyi P, Papachristou GI, et al. Clinical usefulness of scoring systems to predict severe acute pancreatitis: A systematic review and meta-analysis with pre and post-test probability assessment. United European Gastroenterol J. 2023;11(9):825–36. Epub 20230927. doi: 10.1002/ueg2.12464. PubMed PMID: 37755341; PubMed Central PMCID: PMCPMC10637128.

20. Papachristou GI, Muddana V, Yadav D, O’Connell M, Sanders MK, Slivka A, Whitcomb DC. Comparison of BISAP, Ranson’s, APACHE-II, and CTSI scores in predicting organ failure, complications, and mortality in acute pancreatitis. The American journal of gastroenterology. 2010;105(2):435–41; quiz 42. doi: 10.1038/ajg.2009.622.

21. Mounzer R, Langmead CJ, Wu BU, Evans AC, Bishehsari F, Muddana V, et al. Comparison of existing clinical scoring systems to predict persistent organ failure in patients with acute pancreatitis. Gastroenterology. 2012;142(7):1476-. doi: 10.1053/j.gastro.2012.03.005.

22. Sidey-Gibbons JAM, Sidey-Gibbons CJ. Machine learning in medicine: a practical introduction. BMC Med Res Methodol. 2019;19(1):64. Epub 20190319. doi: 10.1186/s12874-019-0681-4. PubMed PMID: 30890124; PubMed Central PMCID: PMCPMC6425557.

23. Zhou Y, Ge YT, Shi XL, Wu KY, Chen WW, Ding YB, et al. Machine learning predictive models for acute pancreatitis: A systematic review. Int J Med Inform. 2022;157:104641. Epub 20211110. doi: 10.1016/j.ijmedinf.2021.104641. PubMed PMID: 34785488.

24. Langmead C, Lee PJ, Paragomi P, Greer P, Stello K, Hart PA, et al. A Novel 5-Cytokine Panel Outperforms Conventional Predictive Markers of Persistent Organ Failure in Acute Pancreatitis. Clinical and translational gastroenterology. 2021;12(5):e00351-e. doi: 10.14309/ctg.0000000000000351.

25. Fei Y, Gao K, Li W-Q. Artificial neural network algorithm model as powerful tool to predict acute lung injury following to severe acute pancreatitis. Pancreatology : official journal of the International Association of Pancreatology (IAP) [et al]. 2018;18(8):892–9. doi: 10.1016/j.pan.2018.09.007.

26. Andaur Navarro CL, Damen JAA, Takada T, Nijman SWJ, Dhiman P, Ma J, et al. Systematic review finds “spin” practices and poor reporting standards in studies on machine learning-based prediction models. J Clin Epidemiol. 2023;158:99–110. Epub 20230405. doi: 10.1016/j.jclinepi.2023.03.024. PubMed PMID: 37024020.

27. Andaur Navarro CL, Damen JAA, van Smeden M, Takada T, Nijman SWJ, Dhiman P, et al. Systematic review identifies the design and methodological conduct of studies on machine learning-based prediction models. J Clin Epidemiol. 2023;154:8–22. Epub 20221125. doi: 10.1016/j.jclinepi.2022.11.015. PubMed PMID: 36436815.

28. Dhiman P, Ma J, Andaur Navarro CL, Speich B, Bullock G, Damen JAA, et al. Overinterpretation of findings in machine learning prediction model studies in oncology: a systematic review. J Clin Epidemiol. 2023;157:120–33. Epub 20230317. doi: 10.1016/j.jclinepi.2023.03.012. PubMed PMID: 36935090.

29. Hassan A, Critelli B, Lahooti I, Lahooti A, Matzko N, Adams JN, et al. Critical appraisal of machine learning prognostic models for acute pancreatitis: protocol for a systematic review. Diagn Progn Res. 2024;8(1):6. Epub 20240402. doi: 10.1186/s41512-024-00169-1. PubMed PMID: 38561864; PubMed Central PMCID: PMCPMC10986113.

30. van Dijk SM, Hallensleben NDL, van Santvoort HC, Fockens P, van Goor H, Bruno MJ, Besselink MG. Acute pancreatitis: recent advances through randomised trials. Gut. 2017;66(11):2024–32. doi: 10.1136/gutjnl-2016-313595.

31. de-Madaria E, Buxbaum JL, Maisonneuve P, García García de Paredes A, Zapater P, Guilabert L, et al. Aggressive or Moderate Fluid Resuscitation in Acute Pancreatitis. The New England journal of medicine. 2022;387(11):989–1000. doi: 10.1056/NEJMoa2202884.

32. Wolbrink DRJ, van de Poll MCG, Termorshuizen F, de Keizer NF, van der Horst ICC, Schnabel R, et al. Trends in Early and Late Mortality in Patients With Severe Acute Pancreatitis Admitted to ICUs: A Nationwide Cohort Study. Crit Care Med. 2022;50(10):1513–21. Epub 20220725. doi: 10.1097/ccm.0000000000005629. PubMed PMID: 35876365.

33. Sorrento C, Shah I, Yakah W, Ahmed A, Tintara S, Kandasamy C, et al. Inpatient Alcohol Cessation Counseling Is Associated With a Lower 30-Day Hospital Readmission in Acute Alcoholic Pancreatitis. J Clin Gastroenterol. 2022;56(9):e313–e7. Epub 20220110. doi: 10.1097/mcg.0000000000001666. PubMed PMID: 34999646.

34. Onnekink AM, Boxhoorn L, Timmerhuis HC, Bac ST, Besselink MG, Boermeester MA, et al. Endoscopic Versus Surgical Step-Up Approach for Infected Necrotizing Pancreatitis (ExTENSION): Long-term Follow-up of a Randomized Trial. Gastroenterology. 2022;163(3):712–22.e14. Epub 20220514. doi: 10.1053/j.gastro.2022.05.015. PubMed PMID: 35580661.

35. Hallensleben ND, Timmerhuis HC, Hollemans RA, Pocornie S, van Grinsven J, van Brunschot S, et al. Optimal timing of cholecystectomy after necrotising biliary pancreatitis. Gut. 2022;71(5):974–82. Epub 20210716. doi: 10.1136/gutjnl-2021-324239. PubMed PMID: 34272261.

36. Sissingh NJ, Groen JV, Koole D, Klok FA, Boekestijn B, Bollen TL, et al. Therapeutic anticoagulation for splanchnic vein thrombosis in acute pancreatitis: A systematic review and meta-analysis. Pancreatology. 2022;22(2):235–43. Epub 20211222. doi: 10.1016/j.pan.2021.12.008. PubMed PMID: 35012902.

37. Schepers NJ, Bakker OJ, Besselink MG, Ahmed Ali U, Bollen TL, Gooszen HG, et al. Impact of characteristics of organ failure and infected necrosis on mortality in necrotising pancreatitis. Gut. 2018.

38. Vela D, Sharp A, Zhang R, Nguyen T, Hoang A, Pianykh OS. Temporal quality degradation in AI models. Scientific Reports. 2022;12(1):11654. doi: 10.1038/s41598-022-15245-z.

39. Geersing GJ, Bouwmeester W, Zuithoff P, Spijker R, Leeflang M, Moons KG. Search filters for finding prognostic and diagnostic prediction studies in Medline to enhance systematic reviews. PLoS One. 2012;7(2):e32844. Epub 20120229. doi: 10.1371/journal.pone.0032844. PubMed PMID: 22393453; PubMed Central PMCID: PMCPMC3290602.

40. Ingui BJ, Rogers MA. Searching for clinical prediction rules in MEDLINE. J Am Med Inform Assoc. 2001;8(4):391–7. doi: 10.1136/jamia.2001.0080391. PubMed PMID: 11418546; PubMed Central PMCID: PMCPMC130084.

41. Moons KGM, de Groot JAH, Bouwmeester W, Vergouwe Y, Mallett S, Altman DG, et al. Critical Appraisal and Data Extraction for Systematic Reviews of Prediction Modelling Studies: The CHARMS Checklist. PLOS Medicine. 2014;11(10):e1001744. doi: 10.1371/journal.pmed.1001744.

42. Wolff RF, Moons KGM, Riley RD, Whiting PF, Westwood M, Collins GS, et al. PROBAST: A Tool to Assess the Risk of Bias and Applicability of Prediction Model Studies. Annals of internal medicine. 2019;170(1):51–8. doi: 10.7326/M18-1376.

43. Collins GS, Moons KGM, Dhiman P, Riley RD, Beam AL, Van Calster B, et al. TRIPOD+AI statement: updated guidance for reporting clinical prediction models that use regression or machine learning methods. BMJ. 2024;385:e078378. doi: 10.1136/bmj-2023-078378.

44. Chen Z, Wang Y, Zhang H, Yin H, Hu C, Huang Z, et al. Deep Learning Models for Severity Prediction of Acute Pancreatitis in the Early Phase From Abdominal Nonenhanced Computed Tomography Images. Pancreas. 2023;52(1):e45–e53. doi: 10.1097/mpa.0000000000002216. PubMed PMID: 37378899.

45. Ding N, Guo C, Li C, Zhou Y, Chai X. An Artificial Neural Networks Model for Early Predicting In-Hospital Mortality in Acute Pancreatitis in MIMIC-III. Biomed Res Int. 2021;2021:6638919. Epub 20210128. doi: 10.1155/2021/6638919. PubMed PMID: 33575333; PubMed Central PMCID: PMCPMC7864739.

46. Hameed MAB, Alamgir Z. Improving mortality prediction in Acute Pancreatitis by machine learning and data augmentation. Comput Biol Med. 2022;150:106077. Epub 20220911. doi: 10.1016/j.compbiomed.2022.106077. PubMed PMID: 36137318.

47. Hong W, Lu Y, Zhou X, Jin S, Pan J, Lin Q, et al. Usefulness of Random Forest Algorithm in Predicting Severe Acute Pancreatitis. Front Cell Infect Microbiol. 2022;12:893294. Epub 20220610. doi: 10.3389/fcimb.2022.893294. PubMed PMID: 35755843; PubMed Central PMCID: PMCPMC9226542.

48. İnce AT, Silahtaroğlu G, Seven G, Koçhan K, Yıldız K, Şentürk H. Early prediction of the severe course, survival, and ICU requirements in acute pancreatitis by artificial intelligence. Pancreatology. 2023;23(2):176–86. Epub 20221230. doi: 10.1016/j.pan.2022.12.005. PubMed PMID: 36610872.

49. Jin X, Ding Z, Li T, Xiong J, Tian G, Liu J. Comparison of MPL-ANN and PLS-DA models for predicting the severity of patients with acute pancreatitis: An exploratory study. Am J Emerg Med. 2021;44:85–91. Epub 20210122. doi: 10.1016/j.ajem.2021.01.044. PubMed PMID: 33582613.

50. Kimita W, Bharmal SH, Ko J, Petrov MS. Identifying endotypes of individuals after an attack of pancreatitis based on unsupervised machine learning of multiplex cytokine profiles. Transl Res. 2023;251:54–62. Epub 20220718. doi: 10.1016/j.trsl.2022.07.001. PubMed PMID: 35863673.

51. Kiss S, Pintér J, Molontay R, Nagy M, Farkas N, Sipos Z, et al. Early prediction of acute necrotizing pancreatitis by artificial intelligence: a prospective cohort-analysis of 2387 cases. Sci Rep. 2022;12(1):7827. Epub 20220512. doi: 10.1038/s41598-022-11517-w. PubMed PMID: 35552440; PubMed Central PMCID: PMCPMC9098474.

52. Kui B, Pintér J, Molontay R, Nagy M, Farkas N, Gede N, et al. EASY-APP: An artificial intelligence model and application for early and easy prediction of severity in acute pancreatitis. Clin Transl Med. 2022;12(6):e842. doi: 10.1002/ctm2.842. PubMed PMID: 35653504; PubMed Central PMCID: PMCPMC9162438.

53. Li JN, Mu D, Zheng SC, Tian W, Wu ZY, Meng J, et al. Machine learning improves prediction of severity and outcomes of acute pancreatitis: a prospective multi-center cohort study. Sci China Life Sci. 2023;66(8):1934–7. Epub 20230516. doi: 10.1007/s11427-022-2333-8. PubMed PMID: 37209250.

54. Liang H, Wang M, Wen Y, Du F, Jiang L, Geng X, et al. Predicting acute pancreatitis severity with enhanced computed tomography scans using convolutional neural networks. Sci Rep. 2023;13(1):17514. Epub 20231016. doi: 10.1038/s41598-023-44828-7. PubMed PMID: 37845380; PubMed Central PMCID: PMCPMC10579320.

55. Luo Z, Shi J, Fang Y, Pei S, Lu Y, Zhang R, et al. Development and evaluation of machine learning models and nomogram for the prediction of severe acute pancreatitis. J Gastroenterol Hepatol. 2023;38(3):468–75. Epub 20230127. doi: 10.1111/jgh.16125. PubMed PMID: 36653317.

56. Luo J, Lan L, Huang S, Zeng X, Xiang Q, Li M, et al. Real-time prediction of organ failures in patients with acute pancreatitis using longitudinal irregular data. J Biomed Inform. 2023;139:104310. Epub 20230210. doi: 10.1016/j.jbi.2023.104310. PubMed PMID: 36773821.

57. Ren W, Zou K, Chen Y, Huang S, Luo B, Jiang J, et al. Application of a Machine Learning Predictive Model for Recurrent Acute Pancreatitis. J Clin Gastroenterol. 2023. Epub 20231103. doi: 10.1097/mcg.0000000000001936. PubMed PMID: 37983784.

58. Shi N, Lan L, Luo J, Zhu P, Ward TRW, Szatmary P, et al. Predicting the Need for Therapeutic Intervention and Mortality in Acute Pancreatitis: A Two-Center International Study Using Machine Learning. J Pers Med. 2022;12(4). Epub 20220411. doi: 10.3390/jpm12040616. PubMed PMID: 35455733; PubMed Central PMCID: PMCPMC9031087.

59. Thapa R, Iqbal Z, Garikipati A, Siefkas A, Hoffman J, Mao Q, Das R. Early prediction of severe acute pancreatitis using machine learning. Pancreatology. 2022;22(1):43–50. Epub 20211016. doi: 10.1016/j.pan.2021.10.003. PubMed PMID: 34690046.

60. Xu F, Chen X, Li C, Liu J, Qiu Q, He M, et al. Prediction of Multiple Organ Failure Complicated by Moderately Severe or Severe Acute Pancreatitis Based on Machine Learning: A Multicenter Cohort Study. Mediators Inflamm. 2021;2021:5525118. Epub 20210503. doi: 10.1155/2021/5525118. PubMed PMID: 34054342; PubMed Central PMCID: PMCPMC8112913.

61. Yan J, Yilin H, Di W, Jie W, Hanyue W, Ya L, Jie P. A nomogram for predicting the risk of mortality in patients with acute pancreatitis and Gram-negative bacilli infection. Front Cell Infect Microbiol. 2022;12:1032375. Epub 20221110. doi: 10.3389/fcimb.2022.1032375. PubMed PMID: 36439207; PubMed Central PMCID: PMCPMC9685314.

62. Yang D, Zhao L, Kang J, Wen C, Li Y, Ren Y, et al. Development and validation of a predictive model for acute kidney injury in patients with moderately severe and severe acute pancreatitis. Clin Exp Nephrol. 2022;26(8):770–87. Epub 20220416. doi: 10.1007/s10157-022-02219-8. PubMed PMID: 35430680.

63. Yang Y, Xiao W, Liu X, Zhang Y, Jin X, Li X. Machine Learning-Assisted Ensemble Analysis for the Prediction of Acute Pancreatitis with Acute Kidney Injury. Int J Gen Med. 2022;15:5061–72. Epub 20220517. doi: 10.2147/ijgm.S361330. PubMed PMID: 35607360; PubMed Central PMCID: PMCPMC9123915.

64. Yang D, Kang J, Li Y, Wen C, Yang S, Ren Y, et al. Development of a predictive nomogram for acute respiratory distress syndrome in patients with acute pancreatitis complicated with acute kidney injury. Ren Fail. 2023;45(2):2251591. Epub 20230919. doi: 10.1080/0886022x.2023.2251591. PubMed PMID: 37724533; PubMed Central PMCID: PMCPMC10512859.

65. Yin M, Zhang R, Zhou Z, Liu L, Gao J, Xu W, et al. Automated Machine Learning for the Early Prediction of the Severity of Acute Pancreatitis in Hospitals. Front Cell Infect Microbiol. 2022;12:886935. Epub 20220610. doi: 10.3389/fcimb.2022.886935. PubMed PMID: 35755847; PubMed Central PMCID: PMCPMC9226483.

66. Yuan L, Ji M, Wang S, Wen X, Huang P, Shen L, Xu J. Machine learning model identifies aggressive acute pancreatitis within 48 h of admission: a large retrospective study. BMC Med Inform Decis Mak. 2022;22(1):312. Epub 20221129. doi: 10.1186/s12911-022-02066-3. PubMed PMID: 36447180; PubMed Central PMCID: PMCPMC9707001.

67. Zhang W, Chang Y, Ding Y, Zhu Y, Zhao Y, Shi R. To Establish an Early Prediction Model for Acute Respiratory Distress Syndrome in Severe Acute Pancreatitis Using Machine Learning Algorithm. J Clin Med. 2023;12(5). Epub 20230221. doi: 10.3390/jcm12051718. PubMed PMID: 36902504; PubMed Central PMCID: PMCPMC10002486.

68. Zhang J, Lv Y, Hou J, Zhang C, Yua X, Wang Y, et al. Machine learning for post-acute pancreatitis diabetes mellitus prediction and personalized treatment recommendations. Sci Rep. 2023;13(1):4857. Epub 20230324. doi: 10.1038/s41598-023-31947-4. PubMed PMID: 36964219; PubMed Central PMCID: PMCPMC10038980.

69. Zhang M, Pang M. Early prediction of acute respiratory distress syndrome complicated by acute pancreatitis based on four machine learning models. Clinics (Sao Paulo). 2023;78:100215. Epub 20230503. doi: 10.1016/j.clinsp.2023.100215. PubMed PMID: 37196588; PubMed Central PMCID: PMCPMC10199163.

70. Zhao Y, Wei J, Xiao B, Wang L, Jiang X, Zhu Y, He W. Early prediction of acute pancreatitis severity based on changes in pancreatic and peripancreatic computed tomography radiomics nomogram. Quant Imaging Med Surg. 2023;13(3):1927–36. Epub 20230201. doi: 10.21037/qims-22-821. PubMed PMID: 36915340; PubMed Central PMCID: PMCPMC10006146.

71. Zhou Y, Han F, Shi XL, Zhang JX, Li GY, Yuan CC, et al. Prediction of the severity of acute pancreatitis using machine learning models. Postgrad Med. 2022;134(7):703–10. Epub 20220712. doi: 10.1080/00325481.2022.2099193. PubMed PMID: 35801388.

72. Zhu C, Zhang S, Zhong H, Gu Z, Kang Y, Pan C, et al. Intra-abdominal infection in acute pancreatitis in eastern China: microbiological features and a prediction model. Ann Transl Med. 2021;9(6):477. doi: 10.21037/atm-21-399. PubMed PMID: 33850874; PubMed Central PMCID: PMCPMC8039642.

73. Arina P, Kaczorek MR, Hofmaenner DA, Pisciotta W, Refinetti P, Singer M, et al. Prediction of Complications and Prognostication in Perioperative Medicine: A Systematic Review and PROBAST Assessment of Machine Learning Tools. Anesthesiology. 2024;140(1):85–101. doi: 10.1097/aln.0000000000004764. PubMed PMID: 37944114.

74. Kakarmath S, Esteva A, Arnaout R, Harvey H, Kumar S, Muse E, et al. Best practices for authors of healthcare-related artificial intelligence manuscripts. NPJ Digit Med. 2020;3:134. Epub 20201016. doi: 10.1038/s41746-020-00336-w. PubMed PMID: 33083569; PubMed Central PMCID: PMCPMC7567805.

75. van der Ploeg T, Austin PC, Steyerberg EW. Modern modelling techniques are data hungry: a simulation study for predicting dichotomous endpoints. BMC Medical Research Methodology. 2014;14(1):137. doi: 10.1186/1471-2288-14-137.

76. Li CL, Jiang M, Pan CQ, Li J, Xu LG. The global, regional, and national burden of acute pancreatitis in 204 countries and territories, 1990-2019. BMC Gastroenterol. 2021;21(1):332. Epub 20210825. doi: 10.1186/s12876-021-01906-2. PubMed PMID: 34433418; PubMed Central PMCID: PMCPMC8390209.

77. Celi LA, Cellini J, Charpignon ML, Dee EC, Dernoncourt F, Eber R, et al. Sources of bias in artificial intelligence that perpetuate healthcare disparities-A global review. PLOS Digit Health. 2022;1(3):e0000022. Epub 20220331. doi: 10.1371/journal.pdig.0000022. PubMed PMID: 36812532; PubMed Central PMCID: PMCPMC9931338.

78. Venema E, Wessler BS, Paulus JK, Salah R, Raman G, Leung LY, et al. Large-scale validation of the prediction model risk of bias assessment Tool (PROBAST) using a short form: high risk of bias models show poorer discrimination. J Clin Epidemiol. 2021;138:32–9. Epub 20210624. doi: 10.1016/j.jclinepi.2021.06.017. PubMed PMID: 34175377.

79. Helmrich I, Mikolić A, Kent DM, Lingsma HF, Wynants L, Steyerberg EW, van Klaveren D. Does poor methodological quality of prediction modeling studies translate to poor model performance? An illustration in traumatic brain injury. Diagn Progn Res. 2022;6(1):8. Epub 20220505. doi: 10.1186/s41512-022-00122-0. PubMed PMID: 35509061; PubMed Central PMCID: PMCPMC9068255.

