## Supplementary material for "A Systematic Review of Machine Learning-based Prognostic Models for Acute Pancreatitis: Towards Improving Methods and Reporting Quality": S1 Figure

Studies from databases/registers **(n = 6480)**

MEDLINE (n = 2060)

Embase (n = 1120)

Unspecified (n = 3300)

**Identification**

Studies included in review **(n = 30)**

Studies excluded **(n = 4643)**

Studies assessed for eligibility **(n = 352)**

Studies sought for retrieval **(n = 352)**

Studies screened **(n = 5007)**

References removed **(n = 1473)**

Duplicates identified by Covidence (n = 1473)

Studies excluded **(n = 322)**

Repeat study (n = 1)

Abstract only (n = 23)

Wrong language (n = 14)

Regression model (n = 223)

Wrong study design (n = 43)

Not Machine Learning paper (n = 9)

Wrong date (n = 6)

Review article (n = 1)

Wrong patient population (n = 2)

**Screening**

**Included**

**Supplementary Figure 1: PRISMA Flow diagram**
