## Supplementary material for "A Systematic Review of Machine Learning-based Prognostic Models for Acute Pancreatitis: Towards Improving Methods and Reporting Quality": S1 Table

**Supplementary Table 1:** Search Strategy in Medline

1. Validat$ OR Predict$.ti. OR Rule$.mp. [mp=title, book title, abstract, original title, name of substance word, subject heading word, floating sub-heading word, keyword heading word, organism supplementary concept word, protocol supplementary concept word, rare disease supplementary concept word, unique identifier, synonyms]
2. (Predict$ AND (Outcome$ OR Risk$ OR Model$)).mp. [mp=title, book title, abstract, original title, name of substance word, subject heading word, floating sub-heading word, keyword heading word, organism supplementary concept word, protocol supplementary concept word, rare disease supplementary concept word, unique identifier, synonyms]
3. ((History OR Variable$ OR Criteria OR Scor$ OR Characteristic$ OR Finding$ OR Factor$) AND (Predict$ OR Model$ OR Decision$ OR Identif$ OR Prognos$)).mp. [mp=title, book title, abstract, original title, name of substance word, subject heading word, floating sub-heading word, keyword heading word, organism supplementary concept word, protocol supplementary concept word, rare disease supplementary concept word, unique identifier, synonyms]
4. (Decision$ AND (Model$ OR Clinical$ OR Logistic Models)).mp. [mp=title, book title, abstract, original title, name of substance word, subject heading word, floating sub-heading word, keyword heading word, organism supplementary concept word, protocol supplementary concept word, rare disease supplementary concept word, unique identifier, synonyms]
5. (Prognostic AND (History OR Variable$ OR Criteria OR Scor$ OR Characteristic$ OR Finding$ OR Factor$ OR Model$)).mp. [mp=title, book title, abstract, original title, name of substance word, subject heading word, floating sub-heading word, keyword heading word, organism supplementary concept word, protocol supplementary concept word, rare disease supplementary concept word, unique identifier, synonyms]
6. (Stratification OR “ROC Curve” OR Discrimination OR Discriminate OR “c-statistic” OR “C statistic” OR “Area under the curve” OR AUC OR Calibration OR Indices OR Algorithm OR Multivariable).mp. [mp=title, book title, abstract, original title, name of substance word, subject heading word, floating sub-heading word, keyword heading word, organism supplementary concept word, protocol supplementary concept word, rare disease supplementary concept word, unique identifier, synonyms]
7. 1 or 2 or 3 or 4 or 5 or 6
8. Exp *Pancreatitis-Associated Proteins/ or exp *Pancreatitis/ or exp *Pancreatitis, Acute Hemorrhagic/ or exp *Pancreatitis, Acute Necrotizing/ or exp *Pancreatitis, Alcoholic/
9. (Acute adj3 pancrea*).tw.
10. 8 or 9
11. 7 and 10
