## Supplementary material for "A Systematic Review of Machine Learning-based Prognostic Models for Acute Pancreatitis: Towards Improving Methods and Reporting Quality": S2 Table

1. exp *Pancreatitis-Associated Proteins/ or exp *Pancreatitis/ or exp *Pancreatitis, Acute Hemorrhagic/ or exp *Pancreatitis, Acute Necrotizing/ or exp *Pancreatitis, Alcoholic/

2. (Acute adj3 pancrea*).ti,ab

3. 1 or 2

4. follow-up.mp.

5. prognos*.tw.

6. ep.fs

7. 4 or 5 or 6

8. 3 and 7

**Supplementary Table 2:** Search Strategy in EMBASE
