## Supplementary material for "A Systematic Review of Machine Learning-based Prognostic Models for Acute Pancreatitis: Towards Improving Methods and Reporting Quality": S3 Table

| Author | Participants: ROB | Participants: Applicability | Predictors: ROB | Predictors: Applicability | Outcomes: ROB | Outcomes: Applicability | Analysis: ROB | Overall: ROB | Overall: Applicability |
| --- | --- | --- | --- | --- | --- | --- | --- | --- | --- |
| Ding | High | Low | High | Low | Low | Low | High | High | Low |
| Jin | Low | Low | High | Low | High | Low | High | High | Low |
| Zhu | Unclear | Low | Low | High | High | Low | High | High | High |
| Langmead | Low | Low | Low | High | Low | Low | High | High | High |
| Xu | High | Low | High | Low | High | Low | High | High | High |
| Thapa | High | High | Unclear | Low | High | High | High | High | High |
| Yang | High | Low | High | High | High | Low | High | High | High |
| Shi | High | Low | High | High | High | Low | High | High | High |
| Kiss | High | Low | Low | Low | High | Low | High | High | Low |
| Yang | High | Low | High | Low | High | Low | High | High | Low |
| Kui | Low | Low | Low | Low | Low | Low | Low | Low | Low |
| Hong | High | Low | Unclear |  | High |  | High | High |  |
| Yin | High | Low | High | High | High | Low | High | High | High |
| Zhou | High | Low | Unclear | Low | Unclear | Low | Unclear | High | Low |
| Kimita | High | Low | Unclear | High | High | Unclear | Unclear | High | Unclear |
| Hameed | High | High | High | Low | Low | Low | High | High | High |
| Yan | High | Low | High | Low | Low | Low | High | High | Low |
| Yuan | High | Unclear | Low | High | High | Low | High | High | High |
| Ince | High | Low | Unclear | Low | High | Low | High | High | Low |
| Luo | High | Low | High | High | High | High | High | High | High |
| Luo | High | High | High | High | High | Low | High | High | High |
| Zhang | High | Low | Low | Low | High | Low | High | High | High |
| Zhao | High | Low | High | High | Low | Low | High | High | Low |
| Zhang | High | Low | Low | Low | Unclear | Low | High | High | Low |
| Zhang | High | Low | High | High | High | Low | High | High | High |
| Li | High | Low | Low | Low | Low | Low | High | High | Low |
| Chen | High | Low | High | High | High | Low | High | High | High |

**Supplementary Table 3: PROBAST rating of models from individual studies**
